# The effect of training and workstation adjustability on teleworker discomfort during the COVID-19 pandemic

**DOI:** 10.1101/2021.10.08.21264708

**Authors:** Megan J. McAllister, Patrick A. Costigan, Joshua P. Davies, Tara L. Diesbourg

**Author notes:** Corresponding Author: Tara Diesbourg Ph.D., CPE, R.Kin_(inactive)_, School of Health Sciences, Oakland University, 433 Meadow Brook Rd., Rochester, MI, 48309.

## Abstract

Advancements in telework have increased occupational flexibility for employees and employers alike. However, while effective telework requires planning, the COVID-19 pandemic required many employees to quickly shift to working from home without making sure the requirements for telework were in place beforehand. This study evaluated the transition to telework on university faculty and staff and investigated the effect of one’s telework setup and ergonomics training on work-related discomfort in the at-home environment. Respondents reported increases in new or worsening pain since working from home of 24% and 51%, respectively, suggesting an immediate need for ergonomic interventions, including workstation evaluations, ergonomic training, and individual ergonomic assessments, for those who work from home.

## 1. Introduction

With concurrent advances in computer power and portability in the last few decades, telework has become a viable option to increase occupational flexibility for employees and employers alike (Montreuil and Lippel, 2003). Researchers studying the benefits of telework and flexible work arrangements report that voluntary telework, employer support, and proper workstation setups outside of the workplace are the keys to its success. The most important of these being that telework is voluntary (Åborg et al., 2002; Beauregard et al., 2019; Jaakson and Kallaste, 2010; Ng, 2010). The benefits of voluntary telework include better work-life balance, increased productivity, and a reduced risk of burnout (Baert et al., 2020). Furthermore, successful teleworkers have a particular set of skills that include the ability to work independently without close supervision and to separate their work life from their personal life (Beauregard et al., 2019). Therefore, employees selected for telework should be based on the nature of the job itself and the qualities of the employee (O’Neill et al., 2009). Finally, part-time telework is best, with some days in the office and other days at home (Åborg et al., 2002; Lundberg and Lindfors, 2002; Ng, 2010; Raišienė et al., 2020).

Unfortunately, early in 2020, these best practice guidelines were abandoned for the sake of employee health and protection from COVID-19. When the COVID-19 pandemic struck, employees were asked to work from home if possible and to maintain a safe distance from others. Overnight the proportion of the US population working from home skyrocketed from approximately 15% to upwards of 60% (Katsabian, 2020). Likewise, the proportion of Canadians working from home increased from 4% to 32% (Mehdi and Morissette, 2021). There was no voluntary opting-in, no self-selection based on job responsibilities, no selection of employees who fit the telework profile, and, more importantly, there was no time to prepare a space in the home to accommodate a full-time teleworking job, all aspects of successful telework as outlined by pre-pandemic research (Åborg et al., 2002; Lundberg and Lindfors, 2002; Ng, 2010; Raišienė et al., 2020).

Under the right conditions even non-voluntary teleworking can be a positive experience. Telework improves work-life balance by allowing individuals to easily move between their professional and family responsibilities (Baert et al., 2020; Buomprisco et al., 2021; de Macêdo et al., 2020; Morilla-Luchena et al., 2021), an effect more pronounced in women (Nguyen and Armoogum, 2021; Raišienė et al., 2020; Ralph et al., 2020). Other benefits of the forced telework imposed during the pandemic include feelings of safety with regards to coronavirus exposure and a decrease in commuting time and its associated risks (Bouziri et al., 2020; Morilla-Luchena et al., 2021). However, this forced shift to telework also had negative effects, including feelings of social isolation, the inability to focus on work amid the demands of the home (especially in households with children and pets), lack of proper workstation equipment and the associated risks of poor workstation setup, longer working hours, increased emotional exhaustion and stress, technological difficulties and issues with accessing work and data remotely, decreased job satisfaction, and the perceived lack of professional advancement (Baert et al., 2020; Buomprisco et al., 2021; Davis et al., 2020; Hadi et al., 2021; Morilla-Luchena et al., 2021; Nguyen and Armoogum, 2021). These negative effects all translate to a reduction in self-reported wellbeing, especially among women (Escudero-Castillo et al., 2021; Ralph et al., 2020). Given the social isolation and fear of the unknown that was related to the global pandemic, many of these factors were unavoidable. However, workstation setup and its associated risks is an area that could have been controlled and possibly overcome had employers and employees been prepared for the sudden telework situation.

An important ergonomic consideration for telework, especially with the inability to prepare beforehand, is that the majority of employees work on laptops. For example, Gerding et al. (2021) recently found that approximately 85% of teleworkers surveyed were working from a laptop at home and only 45% of them had an external monitor. The main problem with laptops is that the screen is coupled to the keyboard, making it impossible to maintain a neutral body position – with one’s head, neck, and spine aligned vertically, shoulders relaxed, and elbows at 90-degrees (Harris and Straker, 2000; Moras and Gamarra, 2007; Price and Dowell, 1998; Sommerich et al., 2002; Straker et al., 1997). The addition of peripheral devices such as an external monitor, keyboard, and mouse to a laptop computer, while not common practice, is often the focus of laptop ergonomics training modules. Studies focused on the efficacy of such training modules show that ergonomics knowledge, posture, body awareness, and work practices while using a laptop improve with specific training (Bowman et al., 2014). Furthermore, the combination of training with appropriate, adjustable office equipment produces better working postures and improved working conditions than either training or adjustable equipment alone (Amick et al., 2003).

Queen’s University has been providing ergonomics consulting services to its office employees for over ten years. These services assess existing risks in an office workstation, adjust workstations to better fit the user, and, where needed, recommend improvements to workstations with additional equipment. These are summarized in customized reports that detail the findings and recommendations. The ergonomic assessment includes an on-site workstation visit and a thorough explanation of the ergonomic hazards identified, how they can be addressed, and educating the client as to why these factors should be considered and their effect on one’s health. Anecdotal reports from university employees suggest that these services reduce musculoskeletal symptoms and improve morale and job satisfaction. Due to COVID-19 restrictions these in-person services were not available as employees moved to a telework situation.

One’s office workstation is likely different than one’s telework workstation due to a difference in equipment and services, such as ergonomic consultations, that might be required to ensure a proper setup. Even those who received workstation evaluations, modifications, a list of recommendations, and education specific to their on-campus office may not have the general knowledge needed to set up their home workstation properly. This could mean that general ergonomic training may be more helpful for workers than focusing on workstation-specific training, as this could help prepare workers for unforeseen circumstances, as seen throughout the COVID-19 pandemic, when ergonomists could not be readily available to help all employees with their home-office workstations.

This study evaluated the transition to telework on university faculty and staff. We investigated the effect of ergonomics training on one’s home office workstation setup and the combined effectiveness of ergonomics training and workstation setup at mitigating work-related discomfort in the at-home work environment. We also evaluated the feasibility of using a survey to assess ergonomic risk factors in a home-based workstation. We hypothesized that workstations that were set up according to ergonomics best practices and allowed for the most adjustability would be associated with lower levels of reported discomfort. We also hypothesized that those workers who had received ergonomic assessments and training on-campus would have improved home-office workstations and would experience less discomfort as a result.

## 2. Materials and Methods

### 2.1. Materials

#### 2.1.1. Survey Development

The survey gathered information similar to that collected during an in-person ergonomic assessment. The first part of the survey included standard demographic questions including age (multiple choice - binned by decade), gender (multiple choice - male, female, other), and occupation (typed word answer). We asked about respondents’ work habits before and after they started working from home and to quantify the time (in hours) spent in different postures (i.e., sitting, standing, walking, kneeling…) as well as the time (in hours) spent at the computer, and the number of times they change positions or change tasks throughout the day. Respondents then compared these values to their work before transitioning to working from home (i.e., increased, decreased, stayed the same). To understand the respondents’ workstation setup, we included a list of typical office workstation devices/elements (e.g., laptop, adjustable chair, number of monitors, mouse, keyboard…) and desk configurations (e.g., bar-height table, standard-height table, sitting on the couch/in bed…) that respondents could choose from. They selected all that applied and added any extra features that were not included.

The next section of the survey focused on perceived discomfort while working. Participants were shown a pain map with 23 body regions highlighted (Fig. 1A) and were asked to select all the areas where they experience discomfort while working from home. For all the regions where they indicated that they experience pain, they quantified that pain on a scale from “No discomfort at all” to “Worst discomfort imaginable”. They were asked to quantify the degree of pain for the time before they started working from home (pre-pandemic) and for how they felt currently.

**Fig. 1:**
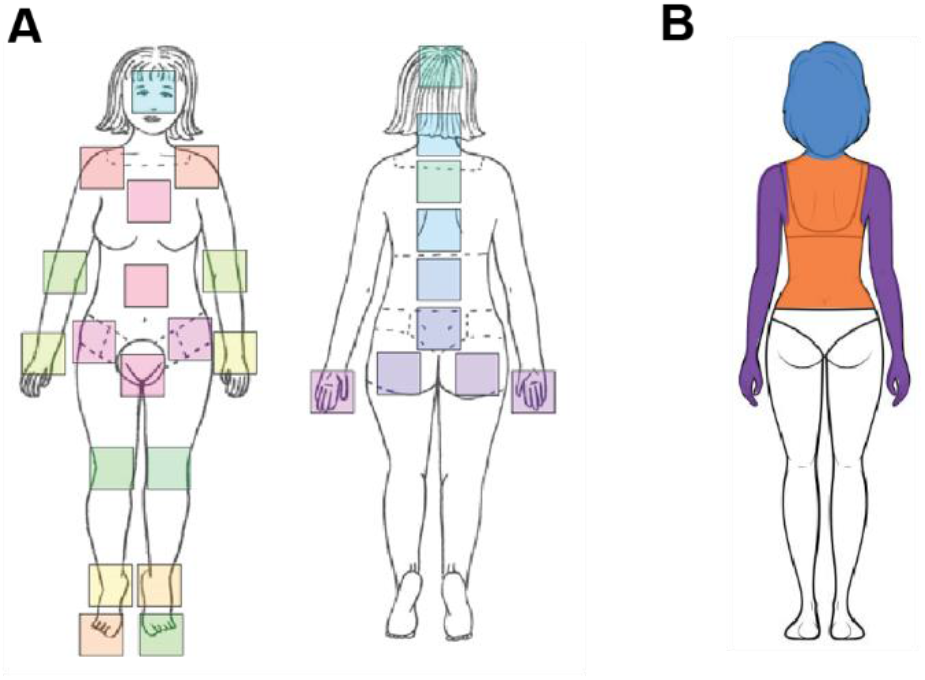
Pain map used in the survey. **A)** The body with 23 body regions highlighted. Respondents selected all the areas where they experience discomfort while working from home. **B)** For analysis, pain areas were defined for three regions: arm (purple), neck (blue) and back (orange). (*One-column fitting image)*

Lastly, we wanted to assess respondents’ ergonomics knowledge to determine if it was related to the quality of their home office workstation setup. They provided information about any ergonomics training they received (e.g., individualized assessment, group assessment, self-directed online search…). Participants who had received an assessment from our ergonomics consulting group were asked for their consent to access their assessment report to identify the workstation elements discussed during their assessment. Respondents could also upload a series of pictures of them at their workstation to provide a better understanding of the quality of their workstation and how well it fit them.

### 2.2. Methods

Prior to beginning the study, the General Research Ethics Board at Queen’s University and the Oakland University Institutional Review Board reviewed and approved all elements of the study, including the recruitment materials and the survey questions.

#### 2.2.1. Participant Recruitment

Study participants were all staff, faculty, and administration at Queen’s University, a large university that provides comprehensive ergonomic consulting services to all employees on campus. We sent recruitment emails to the administrative assistants in all departments, asking them to forward the email to their faculty and staff. All university employees who worked at least part-time at a computer workstation and had been required to work from home during the COVID-19 pandemic were invited to participate in this study. Employees who worked more physical jobs and did not work at computers for at least half of their workday were excluded. Because of the varying levels of support from the department and supervisors, graduate students were not included in the current study. However, graduate students employed by the departments in roles other than teaching and research assistantships were permitted to participate.

All participants who completed the survey and provided their email address were entered into a draw to win a $50 gift card to be used at local businesses. Participants received an additional entry to the draw if they submitted pictures of themselves at their home workstation. Furthermore, participants who submitted workstation pictures were provided with an ergonomics assessment of their workstation to thank them for their support.

#### 2.2.2. Outcome measures

##### 2.2.2.1. Workstation score

As this study was developed in part to evaluate whether assessments of workstation quality could be completed using a survey, a novel scoring system to represent workstation quality was created. In the survey we provided a list of equipment and workstation configurations instructing respondents to “Select all that apply”. The options presented to participants are shown in Table 1.

**Table 1:** Workstation elements contained in the survey to calculate a “Workstation Quality Score” and subsequently, a “Workstation Adjustability Score”.

| Which of the following elements are you using in your "home office" setup? Select all that apply. |  |  |  |
| --- | --- | --- | --- |
| Laptop computer | Three or more external/<br>standalone monitors | Height-adjustable<br>desk | Adjustable office<br>chair |
| Desktop computer | Mouse (not built-in laptop<br>trackpad) | Desk lamp | Separate number pad |
| One external/<br>standalone monitor | Keyboard (not built-in to<br>laptop) | Footrest | Separate sketchpad/<br>trackpad |
| Two external/<br>standalone monitors | Treadmill or pedal<br>ergometer under your desk | Standard-height<br>table | Bar-height table |
| Standing desk | Standard-height computer<br>desk | Sitting on the<br>couch | Sitting in a recliner/<br>easy chair |
| Sitting/laying in bed | With a window directly in<br>front of you | With a window<br>beside you | With a window<br>behind you |

For each element presented in the survey, we assigned a score based on the potential for that piece of equipment to mitigate a person’s work-related discomfort. We assigned a score of zero to a basic computer setup (e.g., computer + mouse + keyboard + one monitor). Beyond that, items that were considered positively influential (would likely reduce discomfort) received a positive score between 1 and 3, where 1 would be minimally influential elements (e.g., desk lamp, separate number pad) and 3 would be maximally influential elements (e.g., height-adjustable desk). Similarly, elements that would negatively contribute to discomfort (would make their discomfort worse) were assigned a negative score from -1 to -3, where -1 would be minimally influential elements (e.g., window behind you) and -3 would be maximally influential (e.g., sitting in bed). Each person’s total workstation score represented their “workstation’s quality”. Upon further reflection, we could not be certain that these elements were used properly and therefore deserved the score that they were assigned. For example, a window behind a worker could create glare on the monitor and contribute to eye fatigue and headaches, however, if the window behind the worker was across a large room, it would not have the same effect. Therefore, we used the workstation elements listed in the survey to calculate a workstation score based on the flexibility/adjustability afforded by the existing equipment. For example, a workstation with a monitor, mouse, and keyboard would be considered more flexible than a workstation with a laptop alone. Likewise, a workstation with an adjustable office chair or a height-adjustable desk would be considered more flexible than a workstation on a dining room table using a non-adjustable chair. As such, a workstation consisting of an office chair, a monitor, a keyboard, and a mouse received a categorical score of 0. A workstation consisting of less equipment than this baseline setup (i.e., working directly on a laptop with no external keyboard or mouse or sitting on a non-adjustable dining room chair) received a categorical score of -1. Conversely, a workstation consisting of more equipment than the baseline setup (i.e., a height-adjustable desk or an additional monitor) received a categorical score of +1.

##### 2.2.2.2. New and worsening pain

We categorized data from the pain map into three key body regions: low back, neck, and arm (Fig. 1B). Respondents reporting discomfort in these three body regions were then further subdivided into groups according to whether that pain was new (did not exist before beginning to work from home), or worsening (pain has worsened since beginning to work from home). We used a clinically relevant threshold of +/-15 to indicate whether pain had increased or decreased (Li et al., 2001; Todd et al., 1996).

##### 2.2.2.3. Ergonomics training

Participants were grouped according to the type of ergonomics training they had received. Anyone who received individualized ergonomics training and assessments from an experienced professional was classified into the “In-person” group, those who completed self-directed online searches were classified into the “Online” group and those having no training were placed into the “No training” group.

#### 2.2.3. Statistical analysis

To determine the relationship between workstation score and ergonomics training, we conducted a bivariate correlation between workstation score (uncategorized; range -3 to +5) and ergonomic training (three categories; in-person, online, no training), with a significance of α = 0.05.

To test the association between workstation score, ergonomic training, and new and worsening pain, we conducted a total of six loglinear analyses (new pain for arm, neck, and back, and worsening pain for arm, neck, and back).

## 3. Results

### 3.1. Telework negatively impacted working conditions

We found that working conditions were worse when employees were working from home. Most respondents (71.0%) reported working their usual 30-40 hours per week (Fig. 2A); however, 64.9% of respondents reported that the amount of time spent at the computer had increased since working from home, while only 3.1% of respondents reported that the amount of time spent at the computer has decreased since working from home (Fig 2B). Moreover, 53.4% of respondents reported changing positions less frequently when working from home, while 17.6% reported no change, and only 29.0% reported changing positions more frequently since working from home (Fig 2C). In addition, most respondents (51.9%) reported no change in the number of times they changed tasks (e.g., going from computer to filing), while 22.1% reported changing tasks more frequently, and 26.0% reported changing tasks less frequently (Fig. 2D).

**Fig. 2:**
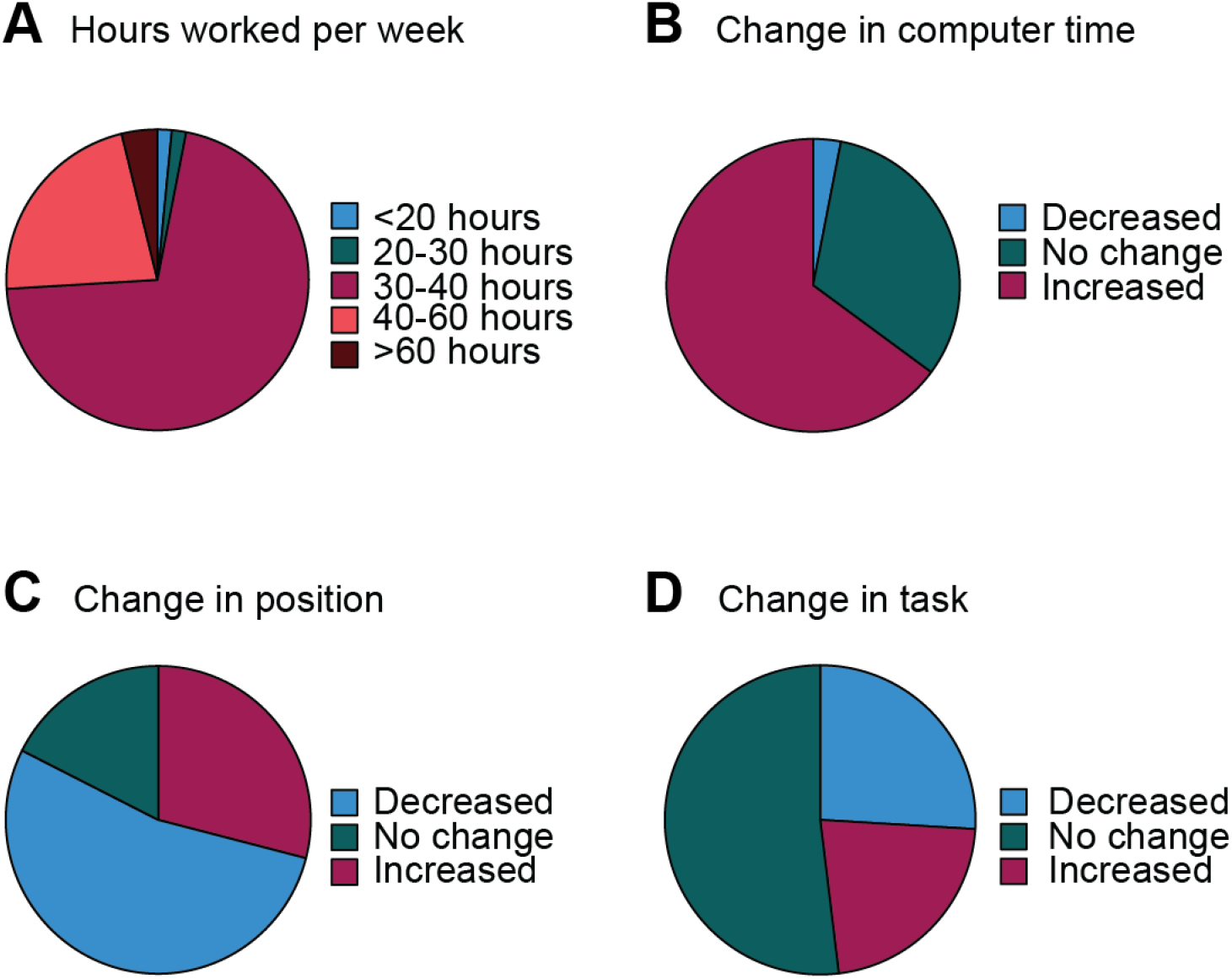
Change in work habits when working from home. A: Reported number of hours worked per week. B: Reported change in time spent working at the computer. C: Reported difference in position changes throughout the workday. D: Reported difference in task changes throughout the workday. (*Two-column fitting image)*

### 3.2. Telework catalyzed new and worsening pain

A disturbing proportion of respondents reported new and worsening pain since working from home (Fig. 3). In total, 51% of respondents reported worsening pain in one or more regions. Of these respondents, 66% reported worsening pain in their arms, 69% reported worsening pain in their neck, and 63% reported worsening pain in their back. Additionally, 24% of respondents reported new pain in at least one region. Of these respondents, 13% reported new pain in their neck, 72% reported new pain in their arms, and 72% reported new pain in their back. Alarmingly, only 7% of respondents reported an improvement in pain since working from home. Of these respondents, 44% reported an improvement in their arms or back, and 67% reported improvement in their neck.

**Fig. 3:**
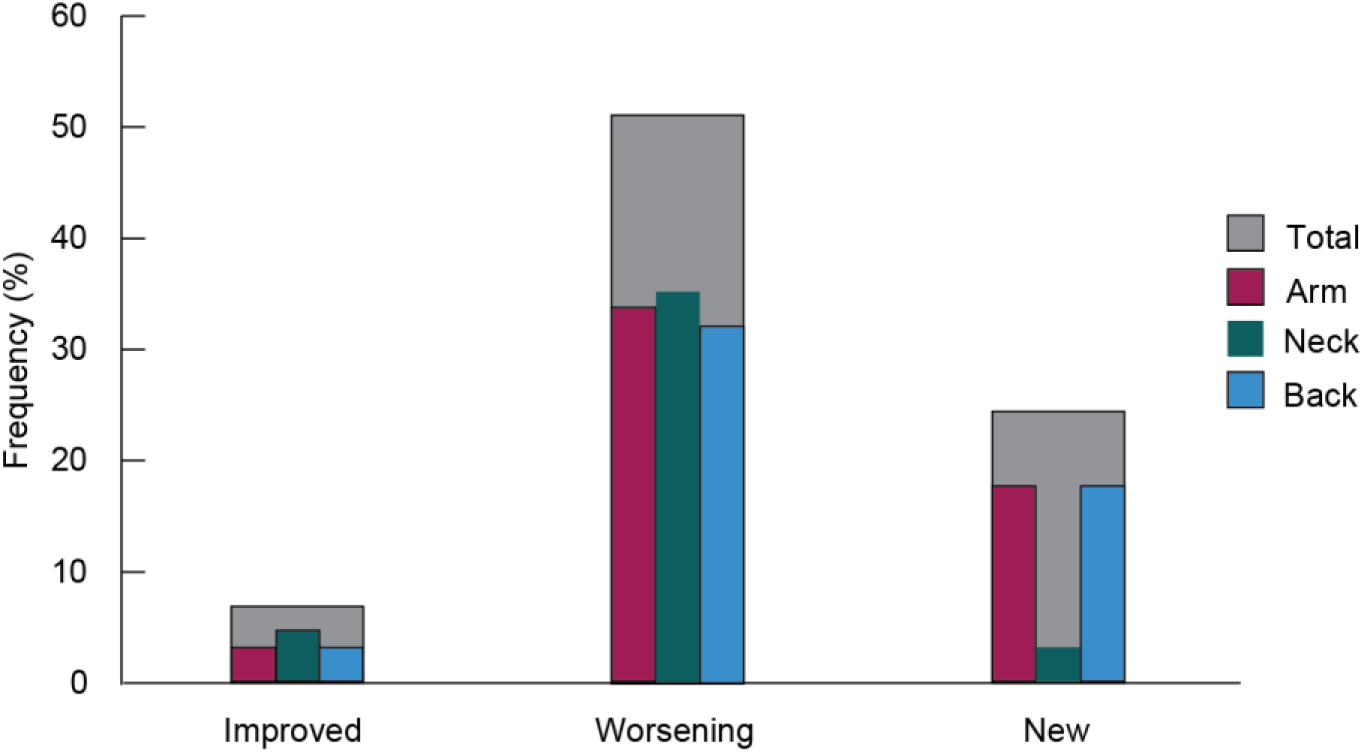
Frequency of improved, worsening, and new pain since working from home. Grey bars represent the total frequency of improved, worsening, and new pain and are further broken down by pain region (red = arm, green = neck, blue = back). *(1*.*5-column fitting image)*

### 3.3. Workstation score and ergonomic training

Most respondents received a score of average (0) or better (≥1) for their workstation equipment. A total of 28 respondents (21%) scored a -1, which is less adequate than a basic setup with a height-adjustable office chair, monitor, keyboard, and mouse. Seventeen respondents (13%) scored a 0, equivalent to a basic setup, and 86 respondents (65%) scored a 1, indicative of an improved workstation setup. As for ergonomic training, 43.5% of respondents reported no training, while 51.1% reported having received an in-person ergonomic assessment and only 5.4% reported online training (e.g., Google search). Surprisingly, the correlation analysis revealed no relationship (r = 0.13, p = 0.16) between training and workstation score using these metrics.

The six three-way loglinear analyses produced a final model that retained only the main effects of workstation score, ergonomic training, and pain (p< .001 for one-way interactions in all loglinear analyses; Appendix I Table S1). There were no significant lower-order interactions between main effects (i.e., workstation score × pain, workstation score × pain, training × pain, and workstation score × training × pain); removing all lower-order interactions does not significantly affect how well the model fits the data (Appendix I Table S1).

## 4. Discussion

The present study evaluated the effect of transitioning to telework on university faculty and staff, the impact of ergonomics training on home office workstation setup, and the combined effectiveness of ergonomics training and workstation setup at mitigating work-related discomfort in the telework environment. In the telework environment participants tended to work at the computer longer and move less. This makes sense as meetings, teaching, and collaborative research move from the office to the screen. There is less movement because there are fewer opportunities to move. With telework you do not have an opportunity to walk to your meeting or classroom. With telework moving will require conscious effort and the knowledge that movement is beneficial in reducing the risk of many musculoskeletal problems.

We found that transitioning to telework had a negative effect on university faculty and staff; only 7% of respondents reported an improvement in their pain/discomfort, while 51% reported worsening pain and 24% reported new pain. Furthermore, we found a positive but insignificant (p=0.08) association between workstation score and ergonomic training, where workstation scores improved as ergonomic training increased from “No training” to “Online” to “In-person”. Lastly, the three-way loglinear analyses between pain, workstation score, and ergonomic training produced a final model that retained only the highest-order interactions (pain × workstation score × ergonomic training; p< .001 for one-way interactions in all loglinear analyses).

There were no significant interactions between ergonomics training, workstation score and either new or increased pain for any body region. That we found no significant two-way or three-way interactions) may be due to the high variability of pain scores. Although we had 134 respondents, the pain ratings were highly variable. Pain is subjective so there is some inherent variability when individuals rate their pain. However, pain scales have been validated and continue to be the most accurate and reliable measure of one’s pain (Karcioglu et al., 2018). We reduced the inter-individual variability by asking participants to rate their pain before and after transitioning to telework and calculating the difference in their pain scores. By using “change in pain” we normalized these scores using each respondent as their own control. However, this normalization required that participants recall their pain from months ago, which poses its own challenges. Although the pain ratings were variable, the reporting of new and worsening pain (24% and 51%, respectively) raise serious concern for the telework environment.

The telework environment varies from home to home and may not be an ideal work environment for some employees. Researchers found that telework had a positive influence on work and personal/family life. However, these findings assume that telework is voluntary and that there is an equivalent workstation setup at home (de Macêdo et al., 2020; Hill et al., 2003; Montreuil and Lippel, 2003). Others report that telework has a negative influence on work-related musculoskeletal disorders (Davis et al., 2020; Escudero-Castillo et al., 2021; Green et al., 2020; Junkin, 2020). Our results agree with these more recent studies, suggesting a negative effect of telework on workstation setup and increased risk of work-related musculoskeletal disorders. In our study, only 7% of respondents reported an improvement in pain since transitioning to telework. Perhaps even more alarming is that 24% of respondents reported new pain when working from home that they did not previously have when working at the office. Furthermore, more than half (51%) of the respondents reported an increase in the severity of their pre-existing work-related pain since transitioning to their telework environment. These alarming proportions suggest a need for ergonomic interventions for those who work from home.

With telework being a potentially permanent solution for many employees (Barrero et al., 2021; Bick et al., 2020), we must develop more effective ergonomic standards and best practices for employees working from home. As suggested by Michael and Smith (2015), telework should be supported by workplace ergonomics programs that provide training, conduct risk assessments, and mitigate potential risks of work-related musculoskeletal disorders. Importantly, this ergonomics program should include two important components: 1) an individualized assessment to uncover risks and provide solutions to mitigate injury risk in the home-office workstation and 2) a training component to provide employees with necessary ergonomics knowledge about work practices and body positioning. Amick et al. (2003) found that workers who received ergonomic training plus a highly adjustable office chair reduced musculoskeletal symptoms over the workday. In comparison, simply adding an adjustable ergonomic office chair while not providing training on its use and setup did not mitigate the musculoskeletal discomfort (Amick et al., 2003). Although the individualized assessments may not seem possible in a telework environment, Blake and Taylor recently validated an approach to provide employees with virtual assessments (2021). Their proposed approach includes three parts: 1) a pre-assessment discomfort survey (much like the one we used in this study), 2) videos of the employee working at their workstation, and 3) a live virtual assessment where the assessor provides recommendations for improving the workstation (Blake and Taylor, 2021).

Based on our results, there is a need to address the lack of telework ergonomics. Although we found a positive trend between ergonomics training and workstation adjustability scores, we did not find any interactions between ergonomics training, workstation adjustability scores, and pain. One reason for this may be that the survey data and the workstation adjustability score did not assess the interaction between the employee and their workstation. A more robust approach would be to develop a framework that considers how well the equipment fits the employee and how they are using the adjustable elements of their workstation.

In summary, employees working in a telework environment should be supported by an ergonomics program that provides adjustable office equipment, necessary ergonomics training, and a virtual assessment to ensure proper workstation setup. Future directions could include the development of a standardized telework package, such that any employee who transitions to teleworking is provided the same set of highly adjustable ergonomic office equipment, the training needed to adjust this equipment, and access to a trained ergonomist who can help address any unique issues faced in setting up the equipment in the home office. Although it would be an added cost for employers in the short-term, implementing this type of program would standardize home-office setups such that discomfort is person-specific, rather than workstation-specific, and can be addressed relatively quickly with minor alterations, thereby improving employee morale and productivity, benefits that employers could use to justify the added cost of implementing such a progressive ergonomics program. However, it is important to note that this system only works if all three elements are present: a sufficiently adjustable workstation, appropriate training, and subject-specific alterations as needed. Without all three elements, the discomfort might persist or even worsen when a worker transitions to a teleworking arrangement.

## Supporting information

Loglinear Analysis Data

## Data Availability

All data produced in the present study are available upon reasonable request to the authors

## 5. Acknowledgments

We would like to thank employees from Queen’s University for their time and participation in our survey.

