## Supplementary material for "The effect of training and workstation adjustability on teleworker discomfort during the COVID-19 pandemic": Loglinear Analysis Data

Table S1: Three-way loglinear analysis between workstation score, ergonomic training, and new/worsening pain for arm, neck, and back regions.

| **Three-way loglinear analysis** | **χ2** | **df** | ***p*-value** |
|---|---|---|---|
| ***Association between workstation score, ergonomic training, and new neck pain*** | ***10.210*** | ***12*** | ***0.598*** |
| New neck pain | 145.817 | 1 | <0.001 |
| Workstation score | 23.092 | 2 | <0.001 |
| Ergonomic training | 62.114 | 2 | <0.001 |
| ***Association between workstation score, ergonomic training, and new arm pain*** | ***11.222*** | ***12*** | ***0.510*** |
| New arm pain | 59.876 | 1 | <0.001 |
| Workstation score | 23.092 | 2 | <0.001 |
| Ergonomic training | 62.114 | 2 | <0.001 |
| ***Association between workstation score, ergonomic training, and new back pain*** | ***12.297*** | ***12*** | ***0.422*** |
| New back pain | 59.876 | 1 | <0.001 |
| Workstation score | 23.092 | 2 | <0.001 |
| Ergonomic training | 62.114 | 2 | <0.001 |
| ***Association between workstation score, ergonomic training, and worsening neck pain*** | ***13.018*** | ***12*** | ***0.368*** |
| Worsening neck pain | 11.789 | 1 | <0.001 |
| Workstation score | 23.092 | 2 | <0.001 |
| Ergonomic training | 62.114 | 2 | <0.001 |
| ***Association between workstation score, ergonomic training, and worsening arm pain*** | ***13.368*** | ***12*** | ***0.343*** |

| | | | |
|---|---|---|---|
| Worsening arm pain | 14.380 | 1 | <0.001 |
| Workstation score | 23.092 | 2 | <0.001 |
| Ergonomic training | 62.114 | 2 | <0.001 |
| ***Association between workstation score, ergonomic training, and worsening back pain*** | ***17.398*** | ***12*** | ***0.135*** |
| Worsening back pain | 17.244 | 1 | <0.001 |
| Workstation score | 23.092 | 2 | <0.001 |
| Ergonomic training | 62.114 | 2 | <0.001 |

1. The three-way loglinear analysis between workstation score, ergonomic training, and *new neck pain* produced a final model that retained the main effects. The likelihood ratio of this was $c^2(12) = 10.210$, $p = 0.598$. This indicates that the lowest-order effects of new neck pain, workstation score, and ergonomic training were significant, $c^2(1) = 145.817$, $p < 0.001$, $c^2(2) = 23.092$, $p < 0.001$, and $c^2(2) = 62.114$, $p < 0.001$, respectively.

2. The three-way loglinear analysis between workstation score, ergonomic training, and *new arm pain* produced a final model that retained the main effects. The likelihood ratio of this was $c^2(12) = 11.222$, $p = 0.510$. This indicates that the lowest-order effects of new arm pain, workstation score, and ergonomic training were significant, $c^2(1) = 59.876$, $p < 0.001$, $c^2(2) = 23.092$, $p < 0.001$, and $c^2(2) = 62.114$, $p < 0.001$, respectively.

3. The three-way loglinear analysis between workstation score, ergonomic training, and *new back pain* produced a final model that retained the main effects. The likelihood ratio of this was $c^2(12) = 12.297$, $p = 0.422$. This indicates that the lowest-order effects of new back pain, workstation score, and ergonomic training were significant, $c^2(1) = 59.876$, $p < 0.001$, $c^2(2) = 23.092$, $p < 0.001$, and $c^2(2) = 62.114$, $p < 0.001$, respectively.

4. The three-way loglinear analysis between workstation score, ergonomic training, and *worsening neck pain* produced a final model that retained the main effects. The likelihood ratio of this was $c^2(12) = 13.018$, $p = 0.368$. This indicates that the lowest-order effects of worsening neck pain, workstation score, and ergonomic training were significant, $c^2(1) = 11.789$, $p = 0.001$, $c^2(2) = 23.092$, $p < 0.001$, and $c^2(2) = 62.114$, $p < 0.001$, respectively.

5. The three-way loglinear analysis between workstation score, ergonomic training, and *worsening arm pain* produced a final model that retained the main effects. The likelihood ratio of this was $c^2(12) = 13.368$, $p = 0.343$. This indicates that the lowest-order effects of

worsening arm pain, workstation score, and ergonomic training were significant, $c^2(1) = 14.380$, $p < 0.001$, $c^2(2) = 23.092$, $p < 0.001$, and $c^2(2) = 62.114$, $p < 0.001$, respectively.

6. The three-way loglinear analysis between workstation score, ergonomic training, and *worsening back pain* produced a final model that retained the main effects. The likelihood ratio of this was $c^2(12) = 17.398$, $p = 0.135$. This indicates that the lowest-order effects of worsening back pain, workstation score, and ergonomic training were significant, $c^2(1) = 17.244$, $p < 0.001$, $c^2(2) = 23.092$, $p < 0.001$, and $c^2(2) = 62.114$, $p < 0.001$, respectively.
